# Genomic monitoring unveil the early detection of the SARS-CoV-2 B.1.351 lineage (20H/501Y.V2) in Brazil

**DOI:** 10.1101/2021.03.30.21254591

**Authors:** Svetoslav Nanev Slavov, José Salvatore Leister Patané, Rafael dos Santos Bezerra, Marta Giovanetti, Vagner Fonseca, Antonio Jorge Martins, Vincent Louis Viala, Evandra Strazza Rodrigues, Elaine Vieira dos Santos, Claudia Renata dos Santos Barros, Elaine Cristina Marqueze, Bibiana Santos, Flavia Aburjaile, Raul Machado Neto, Debora Botequio Moretti, Ricardo Haddad, Rodrigo Tocantins Calado, João Paulo Kitajima, Erika Freitas, David Schlesinger, Luiz Carlos Junior de Alcantara, Maria Carolina Elias, Sandra Coccuzzo Sampaio, Simone Kashima, Dimas Tadeu Covas

**Affiliations:** Blood Center of Ribeirao Preto, Faculty of Medicine of Ribeirao Preto, University of Sao Paulo, Brazil; Butantan Institute, São Paulo, Brazil; Instituto Oswaldo Cruz, FIOCRUZ, Rio de Janeiro, Brazil; Instituto de Ciencias Biologicas, Universidade Federal de Minas Gerais, Belo Horizonte, Minas Gerais, Brazil; Coordenação Geral de Laboratórios de Saúde Pública/Secretaria de Vigilância em Saúde, Ministério da Saúde, (CGLAB/SVS-MS) Brasília, Brazil; KwaZulu-Natal Research Innovation and Sequencing Platform (KRISP), University of KwaZulu-Natal, Durban, South Africa; Faculty of Medicine of Ribeirao Preto, University of Sao Paulo, Brazil; Mendelics Analise Genomica SA, Sao Paulo, Brazil

**Keywords:** SARS-CoV-2, phylogeography, variant of concern, B.1.351 lineage

## Abstract

Sao Paulo State, the most populous area in Brazil, currently experiences a second wave of the COVID-19 pandemic which overwhelmed the healthcare system. Recently, due to the paucity of SARS-CoV-2 complete genome sequences, we established a *Network for Pandemic Alert of Emerging SARS-CoV-2 Variants* to rapidly understand the spread of SARS-CoV-2 and monitor in nearly real-time the circulating SARS-CoV-2 variants into the state. Through full genome analysis of 217 SARS-CoV-2 complete genome sequences obtained from the largest regional health departments we were able to identify the co-circulation of multiple SARS-CoV-2 lineages such as i) B.1.1 (0.92%), ii) B.1.1.1 (0.46%), iii) B.1.1.28 (25.34%), iv) B.1.1.7 (5.99%), v) B.1.566 (1.84%), vi) P.1 (64.05%), and P.2 (0.92%). Further our analysis allowed the detection, for the first time in Brazil of the South African variant of concern (VOC), the B.1.351 (501Y.V2) (0.46%). The identified lineage was characterized by the presence of the following mutations: ORF1ab: T265I, R724K, S1612L, K1655N, K3353R, SGF 3675_F3677del, P4715L, E5585D; Spike: D80A, D215G, L242_L244del, A262D, K417N, E484K, N501Y, D614G, A701V, C1247F; ORF3a: Q57H, S171L, E: P71L; ORF7b: Y10F, N: T205I; ORF14: L52F. Origin of the most recent common ancestor of this genomic variant was inferred to be between middle October to late December 2020. Analysis of generated sequences demonstrated the predominance of the P.1 lineage and allowed the early detection of the South African strain for the first time in Brazil. Our findings highlight the importance to increase active monitoring to ensure the rapid detection of new SARS-CoV-2 variants with a potential impact in pandemic control and vaccination strategies.

## Introduction

The emergence of a new severe acute respiratory syndrome-related coronavirus 2 (SARS-CoV-2) lineage, namely P.1, in the Brazilian city of Manaus [1] and its rapid dissemination despite of the relatively high seroprevalence in the city [2], demonstrated the importance of the SARS-CoV-2 genomic surveillance with the identification of constellations of mutations which may impact viral infectivity, immunological evasion and phenotypic characteristics [1]. Moreover, the importance of these lineages is related to the significant pressure which they exert on the healthcare system of the region due to the high morbidity and mortality leading to an overall collapse of the intensive care units, a situation which occurred when the P1 lineage emerged in the Amazonian city of Manaus.

Apart from data originating from the Brazilian Amazon, the circulating lineages in the Sao Paulo State remain largely unknown. The State incorporates the most populous area of Brazil and is currently experiencing a large second wave of SARS-CoV-2 infections with an estimated total number of cases 2,311,101 up to March, 22, 2021 and a 67,607 deaths with an increase of ∼ 50% of the confirmed SARS-CoV-2 cases. Such a situation gives significant pressure on the healthcare system of the most industrialized state of Brazil. Due to this the *Network for Pandemic Alert of Emerging SARS-CoV-2 Variants* was established in order to implement large-scale genomic surveillance to monitor and characterize the circulating SARS-CoV-2 lineages in the Sao Paulo State. A total of 217 samples were obtained from large cities which constitute the Regional Health Departments of the State by March, 2021. The samples were distributed as follows: 2 obtained from Araçatuba, 64 from Santos, 65 from Campinas, 24 from the Sao Paulo city, 3 from Marilia, 32 from Sorocaba, 26 from Ribeirao Preto and 1 from Taubate. In this study, we also demonstrated for the first time the presence of a B.1.351 lineage in Brazil.

### Study design

We sequenced 217 SARS-CoV-2 samples with RT-qPCR positive results from laboratories which were responsible for SARS-CoV-2 testing in the São Paulo State and were comprising the SARS-CoV-2 Diagnostic Network established by the Butantan Institute. For viral genotyping samples with cycle threshold (Ct) values of up to 30 were only used. Moreover, RNA extraction was performed with the Extracta kit AN viral (Loccus) in an automated extractor (Extracta 32, Loccus) following the manufacturer’s guidelines. SARS-CoV-2 molecular diagnosis was carried out using the kit Gene Finder™ COVID19 Plus RealAmp kit (OSang Healthcare Co. Ltd.) in all laboratories comprising the network which reduces variations related to Ct values.

The SARS-CoV-2 full genome sequencing was performed by Oxford Nanopore Technologies platform using the multiplex PCR amplicon sequencing approach developed by the ARTIC Network (https://www.protocols.io/view/ncov-2019-sequencing-protocol-v3-locost-bh42j8ye). Samples were first reverse transcribed using LunaScript RT SuperMix (New England Biolabs). Amplification was carried out using Q5 Hot Start High-Fidelity 2X Master Mix (New England Biolabs) and a Artik v. 3 primer set, targeting the whole SARS-CoV-2 genome (https://github.com/artic-network/artic-ncov2019/blob/master/primer_schemes/nCoV-2019/V3/nCoV-2019.tsv). Sequencing libraries were prepared using the Oxford Nanopore Ligation Sequencing Kit (SQK-LSK109) and Native Barcoding Expansion kits (NBD104 and EXP-NBD114) following previously published protocol [3]. The libraries were loaded on a R9.4 flow cell (FLO-MIN106) using a MinION MK1B device (ONT).

### Data analysis

Regarding the SARS-CoV-2 genome obtained from the city of Sorocaba, raw files were basecalled using Guppy v4.4.2 (https://nanoporetech.com/nanopore-sequencing-data-analysis) and barcode demultiplexing was performed using qcat v1.1.0 (https://github.com/nanoporetech/qcat). We used Genome Detective [4] and Coronavirus Typing Tool [5] to obtain consensus sequences by de novo assembling. For the remaining genomes, assembly against the SARS-CoV-2 reference (Genbank refseq NC_045512.2) was performed by a pipeline of BWA [6] for mapping, samtools for read indexing and to compile *per* base variation [7], bcftools for variant calling [7], and seqtk for creation of consensus genome [8].

### Phylogenetic and phylodynamic analysis

A representative subset of 3,852 genomes obtained from GISAID [9] was obtained following the Nextstrain guidelines (https://nextstrain.github.io/ncov/customizing-analysis.html). The 217 full length novel genomes were appended to this subset for further analyses. Sequence alignment was performed using MAFFT v7.475 [10] and manually curated to remove artifacts using Aliview [11]. Maximum Likelihood (ML) phylogenetic trees were estimated using FastTree v2.1.10 [12] with local branch support by resampling of site likelihoods 1,000 times in conjunction with the Shimodaira-Hasegawa test.

Beast v1.10.4 [13] was used to infer the date of origin of the Sorocaba haplotype within the B.1.351 clade. A skyride tree prior [14] was employed. Rate prior was incorporated within an uncorrelated lognormal model (UCLN) of rate variation across branches. The parameter ucld.mean (the rate variation prior mean) was set as a uniform [8.99E-4; 1.66E-3] substitutions/site/year following Ghafari et al. (2020) [15]. Two Markov Chain Monte Carlo (MCMC) chains were run in parallel for 100M generations, with convergence assessment checked individually in Tracer v1.7.1 [16], and until all parameters had effective sample sizes (ESS) > 200.

### Mutational patterns analysis

Mutational profile was investigated by using the Nextclade tool (https://clades.nextstrain.org/) to describe substitutions. Subsequently, we compared the set of non-synonymous mutations with the profiles given in the PANGO lineages resource (https://cov-lineages.org/lineages.html) to attribute genomes to lineages.

### Ethical Statement

This study was approved by the Institutional Ethics Committee of the Faculty of Medicine of Ribeirão Preto (Process CAAE: 38975620.1.1001.5440).

## Results

The 217 newly sequenced genomes from the Sao Paulo State classified by Pango lineage demonstrated that the majority of sequences belonged to the P.1 lineage (64.05%) followed by B.1.1.28 (25.34%). Underrepresented lineages were B.1.1 (0.92%), B.1.1.1 (0.46%) and B.1.566 (1.84%). The P.2 Brazilian variant was also detected in 0.92% of the cases. The UK variant of concern (VOC) B.1.1.7 (5.99%) was also detected with a relatively high percentage.

During this genomic surveillance we could identify and characterize for the first time in Brazil a unique isolate classified as B.1.351, which belongs to the South African lineage. This isolate was detected in the southeastern part of Sao Paulo State, and the infected individual reported no recent travel history within and/or outside Brazil. The B.1.351 lineage was characterized by K417N, E484K, N501Y in the spike region and 19 mutations and 2 deletions as follows: ORF1ab: T265I, R724K, S1612L, K1655N, K3353R, SGF 3675_F3677del, P4715L, E5585D; Spike: D80A, D215G, L242_L244del, A262D, D614G, C1247F, ORF3a: Q57H, S171L, E: P71L, ORF7b: Y10F, N: T205I, ORF14: L52F (Table 1). The distribution of SARS-CoV-2 lineages according to Regional Health Departments is shown in Figure 1a.

**Table 1.** Mutational pattern of Brazilian B1.351 variant of concern compared to the South African reference lineage [20] (in bold: common mutations).

| Gene | 501Y.V2 [20] | Brazil B.1.351 VOC |
| --- | --- | --- |
| ORF1ab | **T265I** | **T265I** |
|  | **K1655N** | R724K |
|  | H2799Y | S1612L |
|  | S2900L | **K1655N** |
|  | **K3353R** | **K3353R** |
|  | **SGF 3675-3677del** | **SGF 3675_F3677del** |
|  | D4527Y | P4715L |
|  | T5912I | E5585D |
| Spike | L18F | **D80A** |
|  | **D80A** | **D215G** |
|  | **D215G** | **L242_L244del** |
|  | **L242_244Ldel** | A262D |
|  | R246I | **K417N** |
|  | **K417N** | **E484K** |
|  | **E484K** | **N501Y** |
|  | **N501Y** | D614G |
|  | **A701V** | **A701V** |
|  |  | C1247F |
| ORF3a | **Q57H** | **Q57H** |
|  | **S171L** | **S171L** |
| E | **P71L** | **P71L** |
| ORF7b | - | Y10F |
| N | **T205I** | **T205I** |
| ORF14 | - | L52F |

**Figure 1.**
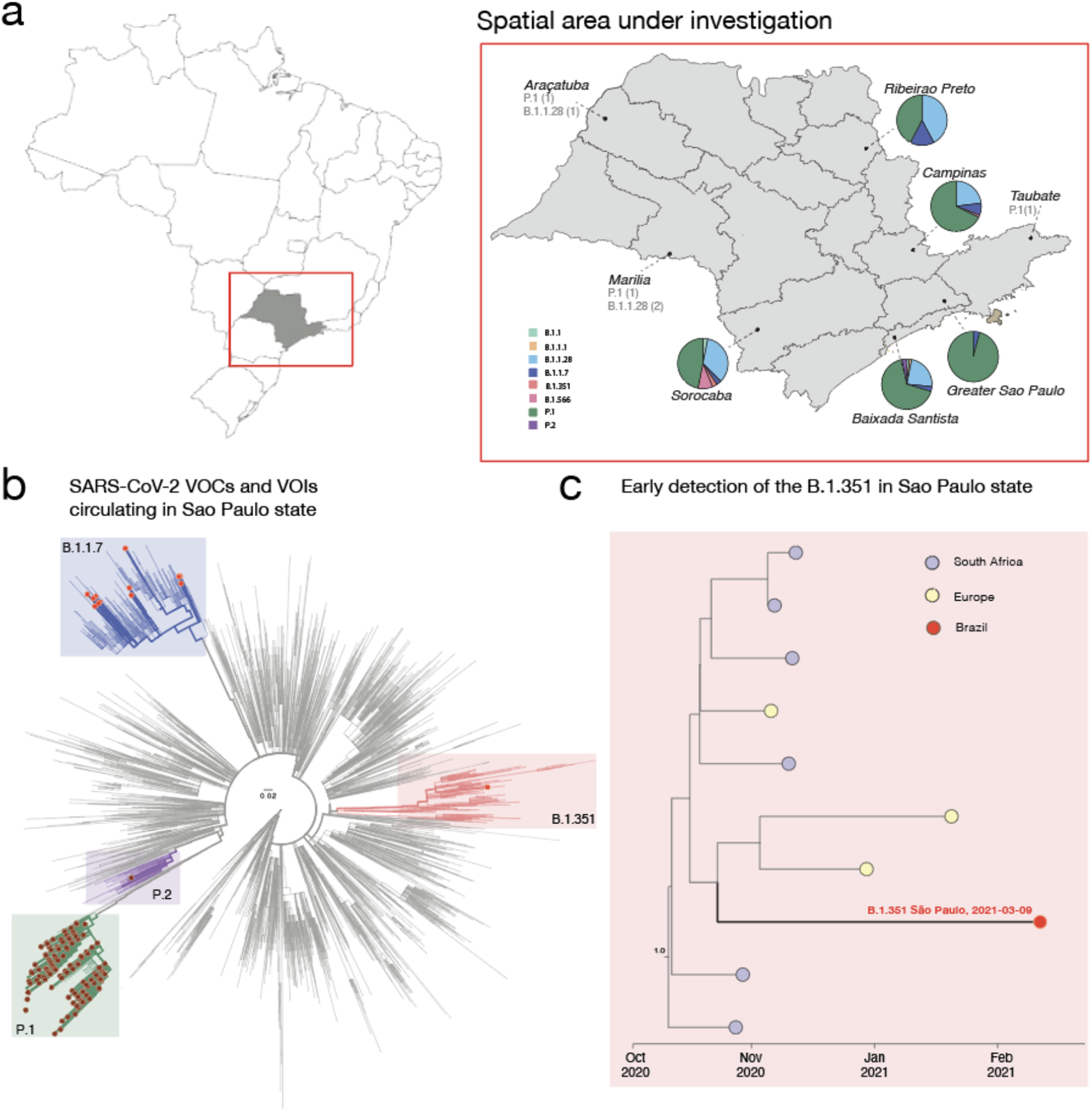
Genomic characterization of SARS-CoV-2 variants circulating in Sao Paulo state. **a)** Map of Brazil and Sao Paulo State (on the right side) showing the frequency of SARS-CoV-2 linhages obtained in this study. **b)** Maximum likelihood (ML) phylogenetic tree including 4,069 genomes from GISAID, including the 217 new isolates obtained in this study plus 3,852 SARS-CoV-2 reference strains collected up to March 21^st^, 2021. **c)** Local subtree showing the closest B.1.351 genomes to the African-like genome from the State of São Paulo (Brazil) after divergence dating.

We then explored the genetic relationship of the newly sequenced SARS-CoV-2 genomes to those of other isolates by phylogenetic inference. Figure 1b highlights the main SARS-CoV-2, VOCs (P.1; B.1.351 ans B.1.1.7) and variants of interest (VOIs) (P.2) circulating into the state. To get more insight regarding the early detection of the SARS-CoV-2 B.1.351 VOC we performed bayesian analysis including all the B.1.315 strain available on GISAID up to March 28^th^, 2021. In Figure 1c, the cluster including the first Brazilian B.1.351 strain is expanded. Although genetic data alone cannot provide evidence of directionality for transmission, our estimates suggest this variant might have reached Brazil through some importation events mediated by returning travellers from European countries, reinforcing how the high connectivity of countries can mediate the introduction of new viral strains. Further, our estimates suggest that the most recent common ancestor of the Brazilian B.1.351 sequenced genome originated between Mid-Oct-2020 and End-Dec-2020 (95% high posterior density).

### Genomic characterization of VOC’s and VOI’s (P.1, P.2 and B.1.1.7)

In order to better characterize the genomes recovered with greater impact in the pandemic, we focus on describing SARS–CoV-2 VOC in more detail. For this we use an analysis where we recover the variations of the genome in relation to the reference strain (NC_045512.2) through the use of bioinformatic pipelines. The UK variant, that is the strain B.1.1.7 (20I / 501Y. V1) is characterized by 14 mutations that define this isolate, 6 of which in spike (ORF1ab: C3267T, C5388A, T6954C, spike: A23063T, C23271A, C23604A, C23709T, T24506G, G24914C, Orf8: C27972T, G28048T, A28111G, nucleocapsid: C28977T). Our strains belonging to this lineage (5.99%) present without exception all these defining mutations. However, a fact that draws attention is the rise of the A23403G (D614G) mutation being present in all samples.

Further we characterized the P.1 (20J / 501Y.V3) VOC containing 15 mutations in its genome, 10 of which were located in the spike region. P.1 defining mutations related to each genomic region were as follows: ORF1ab: S1188L, K1795Q, E5665D, spike: L18F, T20N, P26S, D138Y, R190S, K417T, E484K, N501Y, H655Y, T1027I, Orf8: E92K and nucleocapsid: P80. We additionally observed the D614G mutation in the spike, as well as the mutation G25088T (V1176F). Finally, we characterize the P.2 VOI that was distinguished by five mutations: ORF1ab: C100U, Orf8: C28253U, nucleocapsid: G28628U, G28975U and C29754U and spike: G23012A (E484K).We also highlight the presence of the two mutations - D614G and V1176F - in the spike gene in all analyzed P.1 and P.2 variants of this study.

## Discussion

In this study, we report the dissemination of SARS-CoV-2 variants in the Sao Paulo State. After the notification of the first confirmed case of COVID-19 in Brazil (São Paulo city) in February 2020 [17], São Paulo State experienced two SARS-CoV-2 waves, one between July-August 2020 and the second started in December 2020, which is currently on-going and the State demonstrates uncontrolled number of the COVID-19 cases. To date, the average number of daily confirmed COVID-19 cases increased 1.8-fold, from 100.601 positive cases confirmed by RT-PCR during December, 2020 to 187.810 cases during March, 2021 (unpublished data from The Network of COVID-19 Diagnosis of Sao Paulo State, coordinated by Butantan Institute). Such uncontrolled growth probably reflects VOC spread, as observed during the P.1 emergence in the Amazonian city of Manaus [1,2] and requires a robust SARS-CoV-2 genomic surveillance.

Here, we demonstrated that two main lineages dominate the current SARS-CoV-2 scenario in the State of São Paulo; P.1 and B.1.1.28 which were highly represented (89.39%). However, a dynamic process in the São Paulo State related to substitution of the initial SARS-CoV-2 lineages (B.1.1.28 and B.1.1.33) [18] by a mixture of SARS-CoV-2 VOC including P1, P2, B.1.1.7 and B.1.351 which actually represent 71.42% of circulating SARS-CoV-2 lineages is probably occurring. In this context, the increased transmissibility, which has already been associated with VOC [19] suggest a higher number of positive cases, hospitalizations and mortality observed not only in the Sao Paulo State but in Brazil as general.

The mutational and phylogenetic analysis were consistent with the identification of B1.351 VOC for the first time in Brazil. Based on the molecular, phylogenetic/phylogeographic and epidemiological data, we suggest two potential ways of B1.351 evolution in Brazil. On one hand, the performed phylogenetic analysis clustered the identified B1.351 strain with European and African VOC which suggests that B1.351 presence in Brazil, may have been related to an introduction from travelers originating from one of these countries leading to local transmission. However, a deep analysis of the mutational profile of this strain demonstrates that in comparison with the South African reference, it was carrying three specific mutations in the spike genomic region (A262D, D614G, and C1247F) compared to the reference strain [20]. This suggests that the detected B1.351 VOC might also be considered a convergent strain with local evolution in the Sao Paulo State (or Brazil), once autochthonous transmission has been confirmed. Nevertheless more genomic data surveillance is necessary to evaluate the dissemination and transmission route of the emerging VOC B.1.351 lineage in Brazil.

Other SARS-CoV-2 VOC were also identified, including the UK variant, which shows that the SARS-CoV-2 lineages in the Sao Paulo State are presented as a complex mixture which suggests that more extensive genomic surveillance is urgently needed. In this respect, extensive VOC characterization and collaborative efforts are essential to evaluate the current SARS-CoV-2 scenario not only in the Sao Paulo State but also in Brazil due to the general worsening of the pandemic in a nationwide aspect.

In conclusion, the performed study emphasizes the importance of the SARS-CoV-2 genomic surveillance to monitor the SARS-CoV-2 dissemination in the Brazilian regions and states. Given the vast extension of the country, collaboration of different sequencing networks and combining SARS-CoV-2 genomic data will be of crucial importance to understand in more details the nation-wide Brazilian SARS-CoV-2 pandemic, especially related to VOC emergence and improve the responsiveness to further possible waves and vaccination strategies to SARS-CoV-2.

## Data Availability

The obtained data is being deposited in the GISAID database together with this submission.

## Authors Statements

All authors have no conflicts of interest to declare.

